# Effect of Antibiotic and Steroid Treatment on Leptospirosis Outcomes: A Single-Center Retrospective Cohort Study in the Transcarpathian Region of Ukraine

**DOI:** 10.1101/2024.06.03.24308352

**Authors:** Pavlo Petakh, Valentyn Oksenych, Mykhailo Poliak, Ivan Poliak, Anton Kohutych, Oleksandr Kamyshnyi

## Abstract

Leptospirosis presents a significant health challenge in the Transcarpathian region of Ukraine, with higher incidence rates and mortality compared to national averages. We conducted a retrospective cohort study to investigate the effects of antibiotic and steroid treatments on outcomes in leptospirosis patients. Our analysis of clinical and laboratory data from a single center revealed that dexamethasone showed significant effects on various clinical variables, as did investigated antibiotics. Particularly, direct bilirubin levels emerged as a strong predictor of death. These findings provide valuable insights for clinicians in managing leptospirosis, aiding in outcome prediction and treatment decision-making not only in the Transcarpathian region but globally.

## 1. Introduction

Leptospirosis, a disease primarily found in tropical regions, is a zoonotic illness transmitted through contact with the urine of animals, particularly rats [1]. Its clinical manifestations range from mild to severe, posing life-threatening consequences. Symptoms often mimic those of other infectious diseases, such as influenza [2]. Severe cases, affecting approximately 5% to 15% of individuals, manifest with acute renal failure, acute respiratory distress syndrome, pulmonary issues, hypotension, icterus, and altered mental status [3-7].

In the Transcarpathian region, leptospirosis persists as a prevalent zoonotic disease. Between 2005 and 2015, 420 cases were reported, with an incidence three times higher than the national average. The case fatality rate (CFR) for leptospirosis in Transcarpathia averages 12.5%, exceeding the national level of 9.8% [8-10].

Analysis reveals a significant increase in the notification rate of leptospirosis in Ukraine in 2023. This surge is primarily attributed to the rising incidence of leptospirosis in Transcarpathia, accounting for 150 cases out of the total 433 in Ukraine, and the Ivano-Frankivsk region, contributing 34 cases [11] (Figure 1).

**Figure 1.**
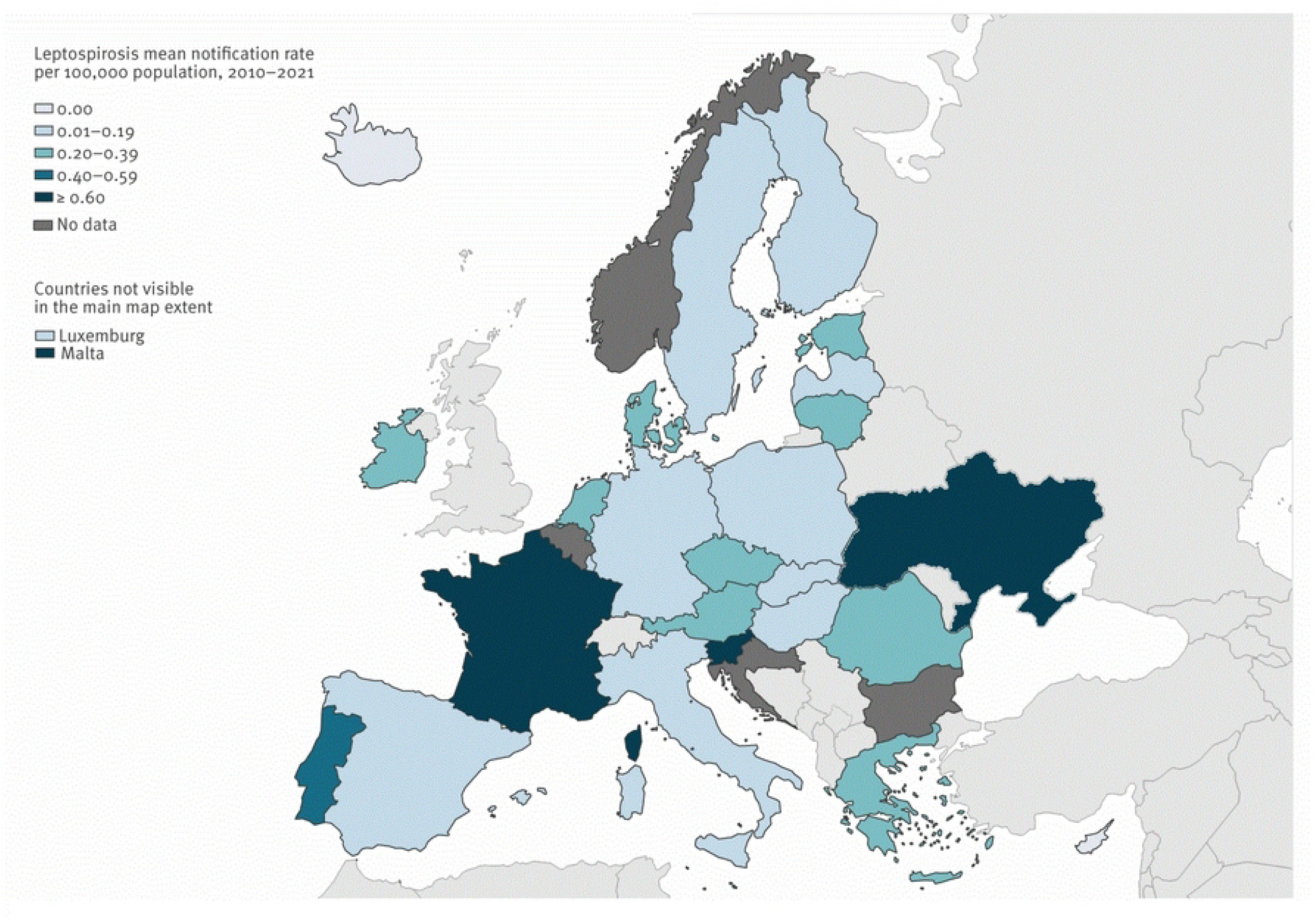
Average annual rate of confirmed leptospirosis cases per 100,000 population, European Union/European Economic Area, 2010–2021. Adapted from ECDC [25]. The average number of leptospirosis cases in Ukraine is one of the highest in Europe, amounting to 0.73 per 100,000 people for the period 2010-2021. For comparison, in neighboring countries, the rate is 0.1 in Poland, 0.15 in Slovakia, and 0.34 in Romania.

Pathogenic Leptospira, responsible for the disease, are excreted in the urine of rats, with humans inadvertently becoming hosts and facing potentially life-threatening consequences [7]. Rats, however, remain immune to fatal infection, serving as natural reservoirs. Most human leptospiral infections are either mild or asymptomatic. Those who do develop the illness typically experience an abrupt onset of symptoms, including fever, rigors, myalgias, and headache [6, 9].

The disease unfolds in two phases [12]. The initial phase involves an acute febrile bacteraemia lasting 2 to 9 days, followed by a period of reduced or no fever and apparent improvement. The second phase, known as the “immune” phase, is characterised by renewed fever and the emergence of complications. Approximately 5%–15% of patients may progress to Weil’s disease, with pulmonary involvement being a notable feature (20%–70%) [7]. Pulmonary complications range from mild cough to severe symptoms like haemoptysis and Acute respiratory distress syndrome (ARDS), the latter carrying a high mortality rate of around 50% [13-15].

The role of corticosteroids in treating severe leptospirosis, especially in addressing pulmonary complications like ARDS, has been explored in a limited number of studies. The argument posits that multi-organ failure in leptospirosis may result from an overactive immune system rather than the direct effects of the pathogen [16]. Therefore, the use of therapeutic doses of steroids is considered to counteract immune activation, potentially reducing mortality and morbidity in severe leptospirosis cases [17]. However, the efficacy of corticosteroids remains uncertain due to the scarcity of studies.

This study aims to investigate the effects of antibiotic and steroid treatment on outcomes in patients diagnosed with leptospirosis within the Transcarpathian Region of Ukraine. Through a retrospective analysis of clinical and laboratory data from a single-center cohort, we aim to assess the impact of these treatments on patient morbidity and mortality rates. Additionally, we aim to explore potential associations between treatment regimens and clinical outcomes, thereby providing significant contributions to the understanding and management of leptospirosis in this geographical area.

## 2. Material and Methods

### 2.1. Study design and diagnostic criteria

We conducted a retrospective single-center cohort study. All medical records were obtained from the Transcarpathian Regional Clinical Infectious Diseases Hospital, Ukraine. Confirmation of diagnosis was determined by the order of the Ministry of Health of Ukraine No. 905 of December 28, 2015. This includes fever or at least two of the following symptoms: chills, headache, myalgia, conjunctival hyperemia, skin and mucous membrane hemorrhages, rash, jaundice, myocarditis, meningitis, kidney failure, respiratory manifestations such as hemoptysis, and laboratory confirmation by Polymerase Chain Reaction (PCR) or MAT (Microscopic Agglutination Test).

#### Inclusion Criteria

Patients included in this study met the following criteria: diagnosed with leptospirosis according to the criteria outlined by the Ministry of Health of Ukraine; with medical records available at the Transcarpathian Regional Clinical Infectious Diseases Hospital; confirmed laboratory diagnosis by PCR or MAT; complete clinical and laboratory data for analysis; aged 18 years and older; and non-pregnant.

#### Exclusion Criteria

Patients were excluded from the study if they met any of the following criteria: incomplete medical records or missing key diagnostic information; incomplete or inconsistent laboratory confirmation results; history of receiving medications that could interfere with the study variables or outcomes; younger than 18 years old; or pregnant.

### 2.2. Patient Stratification and Grouping

We have grouped these patients into different groups for comparing clinical and laboratory data. Firstly, based on sex, males (n=27) and females (n=11). Secondly, based on steroid treatments, the majority received dexamethasone 8 mg per day (n=29), while a few received methylprednisolone 250 mg per day (n=2), and others included patients prescribed prednisolone 1 mg/kg/day or combination steroid types (n=4), with 3 patients not receiving steroids. Next, we grouped the patients based on antibiotic treatment into large groups, i.e. those treated with cephalosporins (n=17), benzylpenicillins (n=8), and others treated with carbapenems or macrolides (n=13). Finally, the patients were also grouped based on outcomes, i.e. 35 survivors and 3 non-survivors.

### 2.3. Statistical analysis

Power analysis was conducted to ascertain the appropriate sample size necessary to detect statistically significant effects or differences in our study variables, while maintaining a desired level of statistical power.

Quantitative variables following a non-normal distribution were described using the median (Me) and lower and upper quartiles (Q1 – Q3). Categorical data were described using absolute and relative frequencies. Comparisons of three or more groups on a quantitative variable with a distribution differing from normality were made using the Kruskal-Wallis test, with Dunn’s criterion and Holm correction applied as a *post hoc* method. The Wilcoxon test was employed for comparing quantitative variables following a non-normal distribution between two matched samples. The Mann-Whitney U-test was used to compare two groups on a quantitative variable with a distribution differing from normality. Frequencies in the analysis of multifield contingency tables were compared using Pearson’s chi-square test.

ROC analysis was utilized to assess the diagnostic performance of quantitative variables in predicting a categorical outcome. The optimal cut-off value of the quantitative variable was estimated using Youden’s J statistic.

Statistical significance was determined at p < 0.05.

## 3. Results

### 3.1. Descriptive analysis and sex-dependent changes in variables

Overall, 38 patients were included in our study, of whom 35 survived and 3 died (7.89%). The socio-demographic and clinical presentations, and laboratory findings of all confirmed patients are shown in Table 1.

**Table 1.** Demographic and Clinical Characteristics of Patients with Leptospirosis in the Transcarpathian Region of Ukraine.

|  | Abs. | % |
| --- | --- | --- |
| **Sex** |  |  |
| Male | 27 | 71.1 |
| Female | 11 | 28.9 |
| **Serogroups** |  |  |
| Hebdomadis | 10 | 26.3 |
| Canicola | 4 | 10.5 |
| Clinical | 3 | 7.9 |
| Others | 2 | 5.3 |
| Pomona | 3 | 7.9 |
| Sejroe | 5 | 13.2 |
| Icterohaemorrhagiae | 3 | 7.9 |
| Cynopteri | 7 | 18.4 |
| Australis | 1 | 2.6 |
| **Outcome** |  |  |
| Survive | 35 | 92.1 |
| Non-survive | 3 | 7.9 |
| **Antibiotic** |  |  |
| Cephalosporins | 17 | 44.7 |
| Benzylpenicillins | 8 | 21.1 |
| Others | 13 | 34.2 |
| **Steroids** |  |  |
| None | 3 | 7.9 |
| Dexamethasone | 29 | 76.3 |
| Methylprednisolone | 2 | 5.3 |
| Others | 4 | 10.5 |

The 38 patients comprised 27 (71.1%) males and 11 (28.9%) females, with a median age of 50.00 years (Table 2). Additionally, we conducted a sex-dependent investigation of clinical and laboratory variables on admission. We found significant changes in Mean Corpuscular Volume (MCV) (p = 0.002), with males having 91.00 (88.50 – 94.30) fL and females having 87.00 (83.50 – 88.50) fL. Males had statistically higher Alanine Aminotransferase (ALT) levels (p = 0.0048) than females, with values of 93.30 (65.20 – 134.15) U/L compared to 61.50 (38.75 – 100.55) U/L, respectively. Similarly, males had higher Aspartate Aminotransferase (AST) levels (p = 0.020) at 81.30 (54.90 – 148.95) U/L compared to females at 47.00 (30.55 – 73.55) U/L.

**Table 2.**
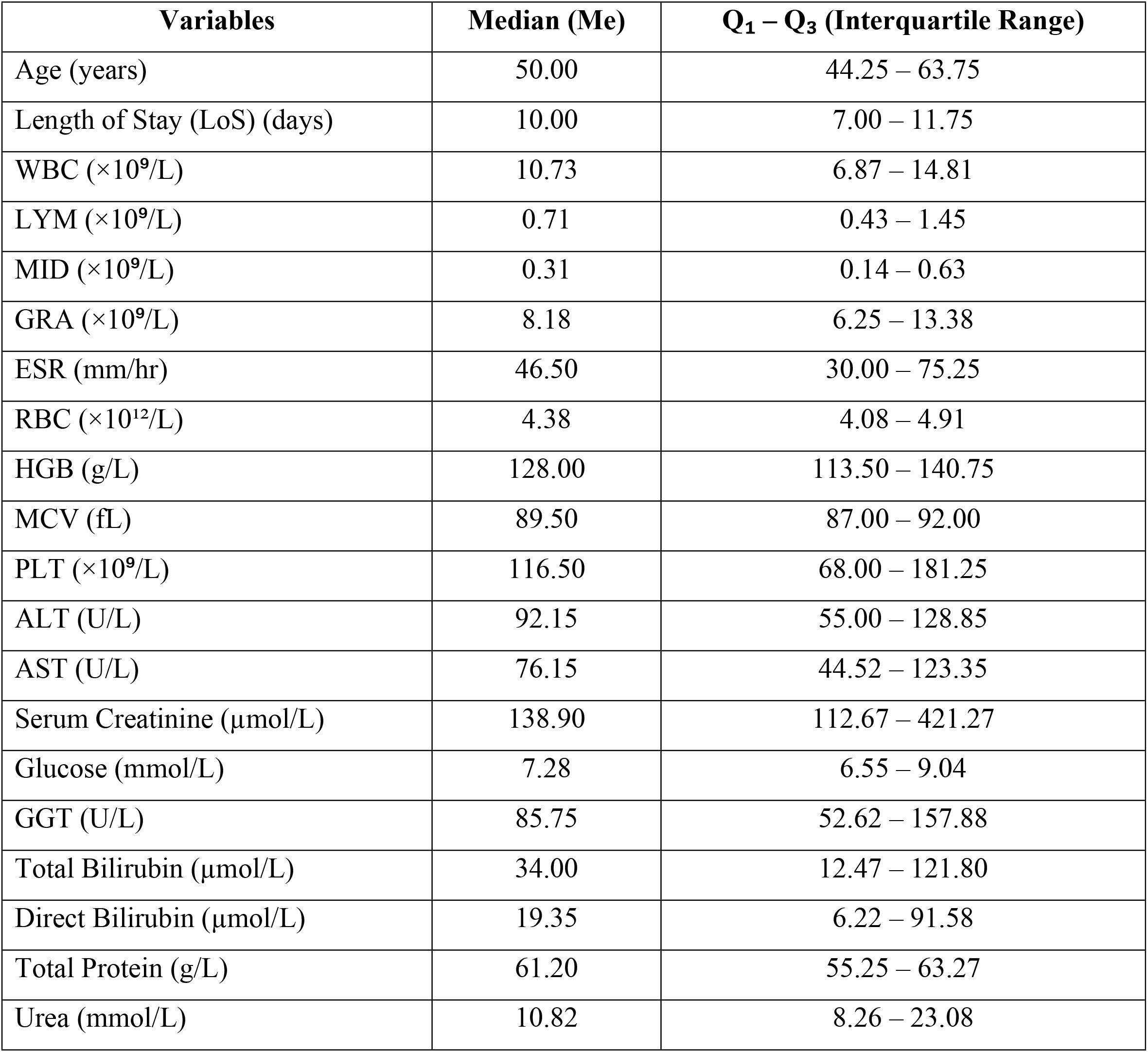
Clinical and Laboratory Findings at Admission for Patients with Leptospirosis.

| Variables | Median (Me) | Q<sub>1</sub> – Q<sub>3</sub> (Interquartile Range) |
| --- | --- | --- |
| Age (years) | 50.00 | 44.25 – 63.75 |
| Length of Stay (LoS) (days) | 10.00 | 7.00 – 11.75 |
| WBC ($\times 10^9/L$) | 10.73 | 6.87 – 14.81 |
| LYM ($\times 10^9/L$) | 0.71 | 0.43 – 1.45 |
| MID ($\times 10^9/L$) | 0.31 | 0.14 – 0.63 |
| GRA ($\times 10^9/L$) | 8.18 | 6.25 – 13.38 |
| ESR (mm/hr) | 46.50 | 30.00 – 75.25 |
| RBC ($\times 10^{12}/L$) | 4.38 | 4.08 – 4.91 |
| HGB (g/L) | 128.00 | 113.50 – 140.75 |
| MCV (fL) | 89.50 | 87.00 – 92.00 |
| PLT ($\times 10^9/L$) | 116.50 | 68.00 – 181.25 |
| ALT (U/L) | 92.15 | 55.00 – 128.85 |
| AST (U/L) | 76.15 | 44.52 – 123.35 |
| Serum Creatinine ($\mu\text{mol}/L$) | 138.90 | 112.67 – 421.27 |
| Glucose (mmol/L) | 7.28 | 6.55 – 9.04 |
| GGT (U/L) | 85.75 | 52.62 – 157.88 |
| Total Bilirubin ($\mu\text{mol}/L$) | 34.00 | 12.47 – 121.80 |
| Direct Bilirubin ($\mu\text{mol}/L$) | 19.35 | 6.22 – 91.58 |
| Total Protein (g/L) | 61.20 | 55.25 – 63.27 |
| Urea (mmol/L) | 10.82 | 8.26 – 23.08 |

### 3.2. Effect of steroid type on clinical and laboratory findings

Most patients took dexamethasone, comprising 29 out of 38 patients (76.3%). Additionally, only 2 patients (5.3%) received methylprednisolone. Four patients were administered other types of steroids, while three patients did not receive any steroid treatment during their hospitalization.

When we compared laboratory findings before admission and after discharge, we found statistically significant changes only in the group that took dexamethasone. The statistically significant changes were observed in Lymphocytes (LYM) levels (on admission: 0.79 ×10^9^/L (0.44 – 1.45), on discharge: 2.30 ×10^9^/L (1.59 – 2.99), p < 0.001), Mid-sized Cells (MID) (on admission:0.30 ×10^9^/L (0.14 – 0.47), on discharge: 0.71 ×10^9^/L (0.56 – 0.81), p = 0.002), Granulocytes (GRA)levels (on admission: 8.18 ×10^9^/L (6.25 – 12.16), on discharge: 6.28 ×10^9^/L (5.00 – 7.87), p = 0.033),Red Blood Cells (RBC) (on admission: 4.64 ×10^12^/L (4.17 – 4.91), on discharge: 4.30 ×10^12^/L (3.63– 4.66), p = 0.004), MCV (on admission: 90.00 fL (87.00 – 92.00), on discharge: 93.00 fL (89.00 – 95.00), p = 0.004), Platelets (PLT) levels (on admission: 123.00 ×10^9^/L (68.00 – 182.00), on discharge: 320.00 ×10^9^/L (204.00 – 350.00), p < 0.001), AST (on admission: 72.30 U/L (37.30 – 122.00), on discharge: 41.50 U/L (30.50 – 58.10), p = 0.016), Direct bilirubin (on admission: 14.40 μmol/L (5.68 – 68.70), on discharge: 10.50 μmol/L (5.70 – 21.20), p = 0.026), and Urea levels (on admission: 9.63 mmol/L (8.22 – 23.02), on discharge: 7.80 mmol/L (6.65 – 8.63), p = 0.005). All other laboratory parameters such as White Blood Cells (WBC) (×10^9^/L), Erythrocyte Sedimentation Rate (ESR) (mm/hr), Hemoglobin (HGB) (g/L), ALT (U/L), Serum Creatinine (μmol/L), glucose (mmol/L), Gamma-Glutamyl Transferase (GGT) (U/L), Total Bilirubin (μmol/L), Total Protein (g/L) did not show significant differences between admission and discharge (p > 0.05) (Figure 2).

**Figure 2.**
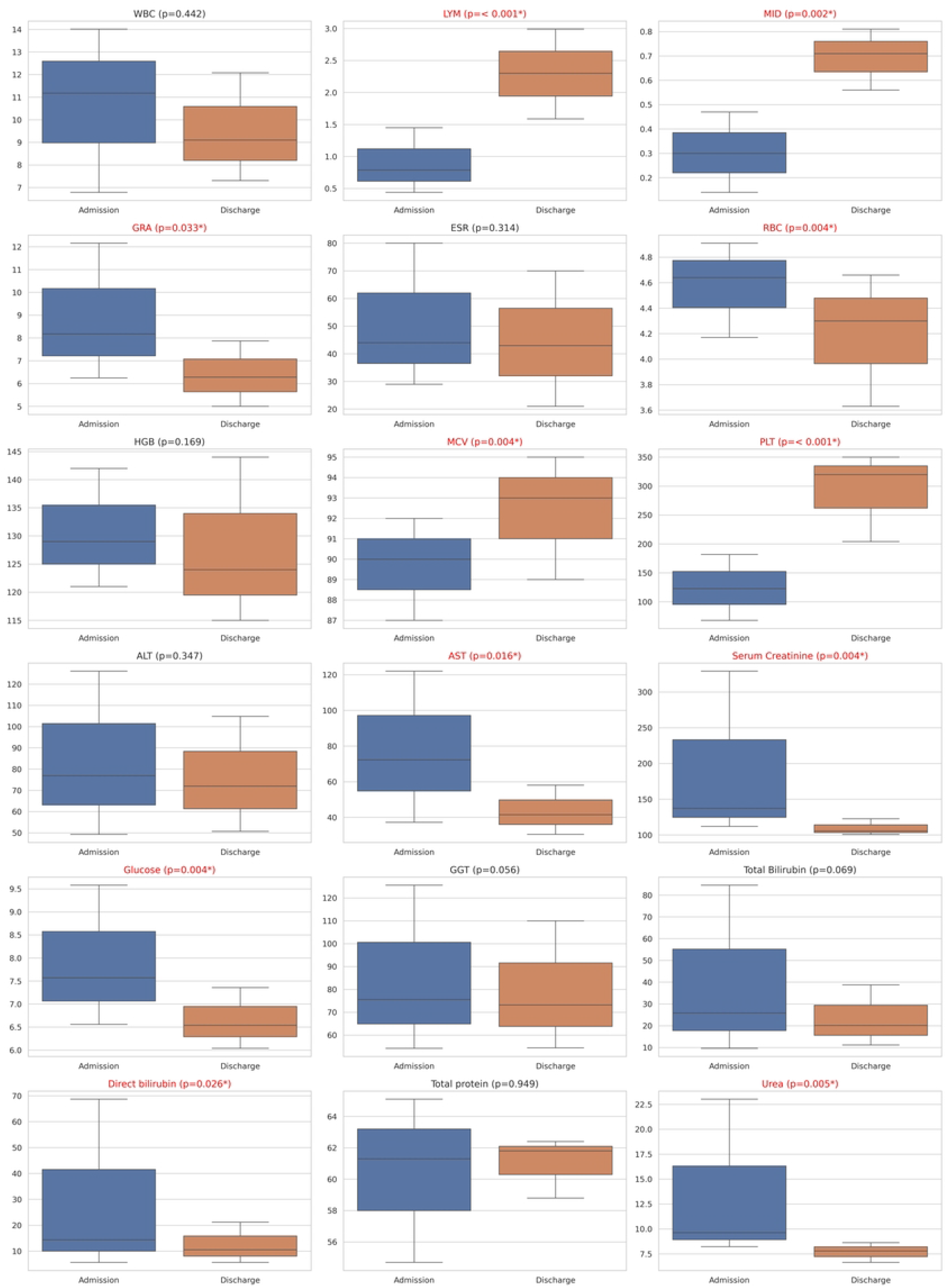
Facet box plots of laboratory findings on admission and discharge in patients who took dexamethasone Statistically significant changes were observed in multiple parameters including LYM, MID, GRA, RBC, MCV, PLT, AST, Direct bilirubin, and Urea levels.

### 3.3. Effect of antibiotic treatment on clinical and laboratory findings

We divided patients into 3 groups, i.e. those who were prescribed cephalosporins - 17 patients, those who were prescribed benzylpenicillin - 8 patients, and a group receiving alternative antimicrobial therapy - 13 patients. We compared the differences in the same indicators as when comparing the effects of steroids.

When comparing laboratory findings before admission and after discharge, we observed statistically significant changes in the LYM (p=0.002), MID (p=0.027), MCV (p=0.006), PLT (p=0.003), AST (p=0.015), glucose (p=0.045), Total Bilirubin (p=0.009), Direct bilirubin (p=0.015), and Urea (p=0.013) for the group treated with cephalosporins. Patients treated with benzylpenicillins exhibited statistically significant changes in LYM (p=0.039), MID (p=0.028), RBC (p=0.039), HGB (p=0.028), and PLT (p=0.039). Patients receiving alternative antimicrobial therapy experienced significant changes in LYM (p=0.033), RBC (p=0.008), MCV (p=0.033), PLT (p=< 0.001), Serum Creatinine (p=< 0.001), Direct bilirubin (p=0.048), and Urea (p=0.010) (Table 3). When we compared the effect of antibiotics and steroids on the length of stay (LoS) and outcomes, we did not find any significant differences across the groups (Figure 3).

**Table 3.**
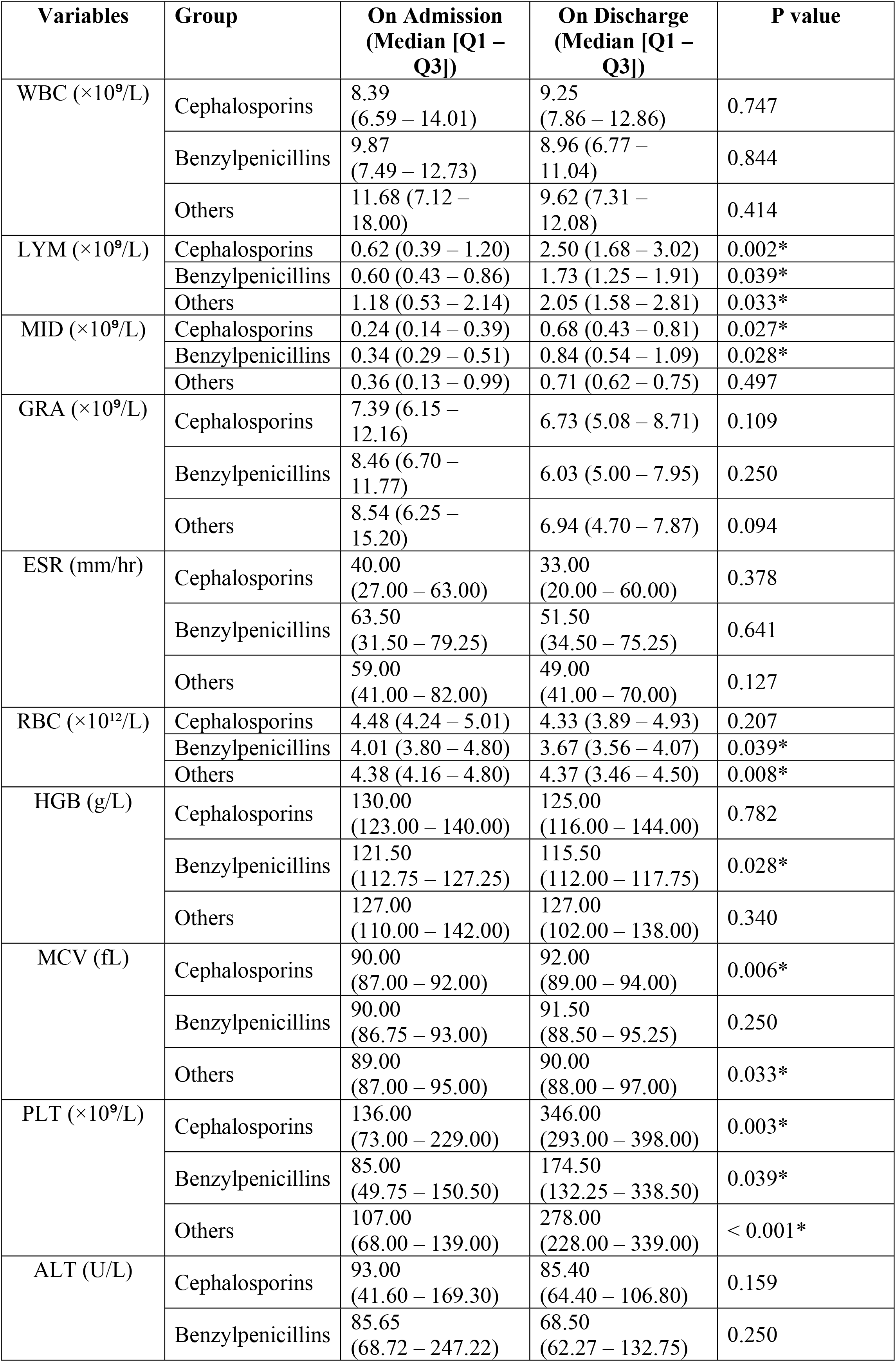
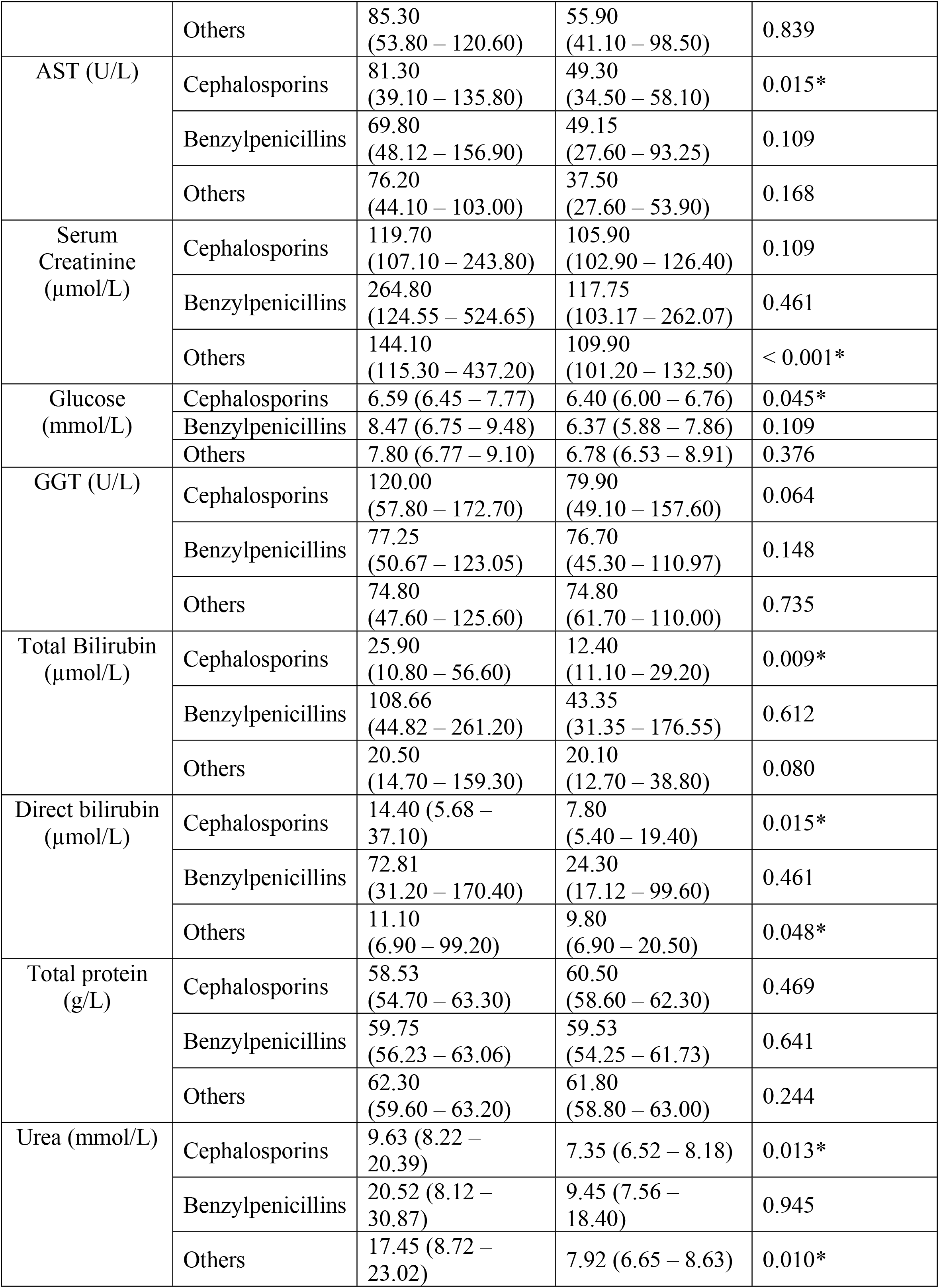
Effect of Antibiotics on Laboratory Findings.

| Variables | Group | On Admission<br>(Median [Q1 – Q3]) | On Discharge<br>(Median [Q1 – Q3]) | P value |
| --- | --- | --- | --- | --- |
| WBC ($\times 10^9/L$) | Cephalosporins | 8.39<br>(6.59 – 14.01) | 9.25<br>(7.86 – 12.86) | 0.747 |
|  | Benzylpenicillins | 9.87<br>(7.49 – 12.73) | 8.96 (6.77 – 11.04) | 0.844 |
|  | Others | 11.68 (7.12 – 18.00) | 9.62 (7.31 – 12.08) | 0.414 |
| LYM ($\times 10^9/L$) | Cephalosporins | 0.62 (0.39 – 1.20) | 2.50 (1.68 – 3.02) | 0.002* |
|  | Benzylpenicillins | 0.60 (0.43 – 0.86) | 1.73 (1.25 – 1.91) | 0.039* |
|  | Others | 1.18 (0.53 – 2.14) | 2.05 (1.58 – 2.81) | 0.033* |
| MID ($\times 10^9/L$) | Cephalosporins | 0.24 (0.14 – 0.39) | 0.68 (0.43 – 0.81) | 0.027* |
|  | Benzylpenicillins | 0.34 (0.29 – 0.51) | 0.84 (0.54 – 1.09) | 0.028* |
|  | Others | 0.36 (0.13 – 0.99) | 0.71 (0.62 – 0.75) | 0.497 |
| GRA ($\times 10^9/L$) | Cephalosporins | 7.39 (6.15 – 12.16) | 6.73 (5.08 – 8.71) | 0.109 |
|  | Benzylpenicillins | 8.46 (6.70 – 11.77) | 6.03 (5.00 – 7.95) | 0.250 |
|  | Others | 8.54 (6.25 – 15.20) | 6.94 (4.70 – 7.87) | 0.094 |
| ESR (mm/hr) | Cephalosporins | 40.00<br>(27.00 – 63.00) | 33.00<br>(20.00 – 60.00) | 0.378 |
|  | Benzylpenicillins | 63.50<br>(31.50 – 79.25) | 51.50<br>(34.50 – 75.25) | 0.641 |
|  | Others | 59.00<br>(41.00 – 82.00) | 49.00<br>(41.00 – 70.00) | 0.127 |
| RBC ($\times 10^{12}/L$) | Cephalosporins | 4.48 (4.24 – 5.01) | 4.33 (3.89 – 4.93) | 0.207 |
|  | Benzylpenicillins | 4.01 (3.80 – 4.80) | 3.67 (3.56 – 4.07) | 0.039* |
|  | Others | 4.38 (4.16 – 4.80) | 4.37 (3.46 – 4.50) | 0.008* |
| HGB (g/L) | Cephalosporins | 130.00<br>(123.00 – 140.00) | 125.00<br>(116.00 – 144.00) | 0.782 |
|  | Benzylpenicillins | 121.50<br>(112.75 – 127.25) | 115.50<br>(112.00 – 117.75) | 0.028* |
|  | Others | 127.00<br>(110.00 – 142.00) | 127.00<br>(102.00 – 138.00) | 0.340 |
| MCV (fL) | Cephalosporins | 90.00<br>(87.00 – 92.00) | 92.00<br>(89.00 – 94.00) | 0.006* |
|  | Benzylpenicillins | 90.00<br>(86.75 – 93.00) | 91.50<br>(88.50 – 95.25) | 0.250 |
|  | Others | 89.00<br>(87.00 – 95.00) | 90.00<br>(88.00 – 97.00) | 0.033* |
| PLT ($\times 10^9/L$) | Cephalosporins | 136.00<br>(73.00 – 229.00) | 346.00<br>(293.00 – 398.00) | 0.003* |
|  | Benzylpenicillins | 85.00<br>(49.75 – 150.50) | 174.50<br>(132.25 – 338.50) | 0.039* |
|  | Others | 107.00<br>(68.00 – 139.00) | 278.00<br>(228.00 – 339.00) | < 0.001* |
| ALT (U/L) | Cephalosporins | 93.00<br>(41.60 – 169.30) | 85.40<br>(64.40 – 106.80) | 0.159 |
|  | Benzylpenicillins | 85.65<br>(68.72 – 247.22) | 68.50<br>(62.27 – 132.75) | 0.250 |
|  | Others | 85.30<br>(53.80 – 120.60) | 55.90<br>(41.10 – 98.50) | 0.839 |
| AST (U/L) | Cephalosporins | 81.30<br>(39.10 – 135.80) | 49.30<br>(34.50 – 58.10) | 0.015* |
|  | Benzylpenicillins | 69.80<br>(48.12 – 156.90) | 49.15<br>(27.60 – 93.25) | 0.109 |
|  | Others | 76.20<br>(44.10 – 103.00) | 37.50<br>(27.60 – 53.90) | 0.168 |
| Serum Creatinine (μmol/L) | Cephalosporins | 119.70<br>(107.10 – 243.80) | 105.90<br>(102.90 – 126.40) | 0.109 |
|  | Benzylpenicillins | 264.80<br>(124.55 – 524.65) | 117.75<br>(103.17 – 262.07) | 0.461 |
|  | Others | 144.10<br>(115.30 – 437.20) | 109.90<br>(101.20 – 132.50) | < 0.001* |
| Glucose (mmol/L) | Cephalosporins | 6.59 (6.45 – 7.77) | 6.40 (6.00 – 6.76) | 0.045* |
|  | Benzylpenicillins | 8.47 (6.75 – 9.48) | 6.37 (5.88 – 7.86) | 0.109 |
|  | Others | 7.80 (6.77 – 9.10) | 6.78 (6.53 – 8.91) | 0.376 |
| GGT (U/L) | Cephalosporins | 120.00<br>(57.80 – 172.70) | 79.90<br>(49.10 – 157.60) | 0.064 |
|  | Benzylpenicillins | 77.25<br>(50.67 – 123.05) | 76.70<br>(45.30 – 110.97) | 0.148 |
|  | Others | 74.80<br>(47.60 – 125.60) | 74.80<br>(61.70 – 110.00) | 0.735 |
| Total Bilirubin (μmol/L) | Cephalosporins | 25.90<br>(10.80 – 56.60) | 12.40<br>(11.10 – 29.20) | 0.009* |
|  | Benzylpenicillins | 108.66<br>(44.82 – 261.20) | 43.35<br>(31.35 – 176.55) | 0.612 |
|  | Others | 20.50<br>(14.70 – 159.30) | 20.10<br>(12.70 – 38.80) | 0.080 |
| Direct bilirubin (μmol/L) | Cephalosporins | 14.40 (5.68 – 37.10) | 7.80<br>(5.40 – 19.40) | 0.015* |
|  | Benzylpenicillins | 72.81<br>(31.20 – 170.40) | 24.30<br>(17.12 – 99.60) | 0.461 |
|  | Others | 11.10<br>(6.90 – 99.20) | 9.80<br>(6.90 – 20.50) | 0.048* |
| Total protein (g/L) | Cephalosporins | 58.53<br>(54.70 – 63.30) | 60.50<br>(58.60 – 62.30) | 0.469 |
|  | Benzylpenicillins | 59.75<br>(56.23 – 63.06) | 59.53<br>(54.25 – 61.73) | 0.641 |
|  | Others | 62.30<br>(59.60 – 63.20) | 61.80<br>(58.80 – 63.00) | 0.244 |
| Urea (mmol/L) | Cephalosporins | 9.63 (8.22 – 20.39) | 7.35 (6.52 – 8.18) | 0.013* |
|  | Benzylpenicillins | 20.52 (8.12 – 30.87) | 9.45 (7.56 – 18.40) | 0.945 |
|  | Others | 17.45 (8.72 – 23.02) | 7.92 (6.65 – 8.63) | 0.010* |

**Figure 3.**
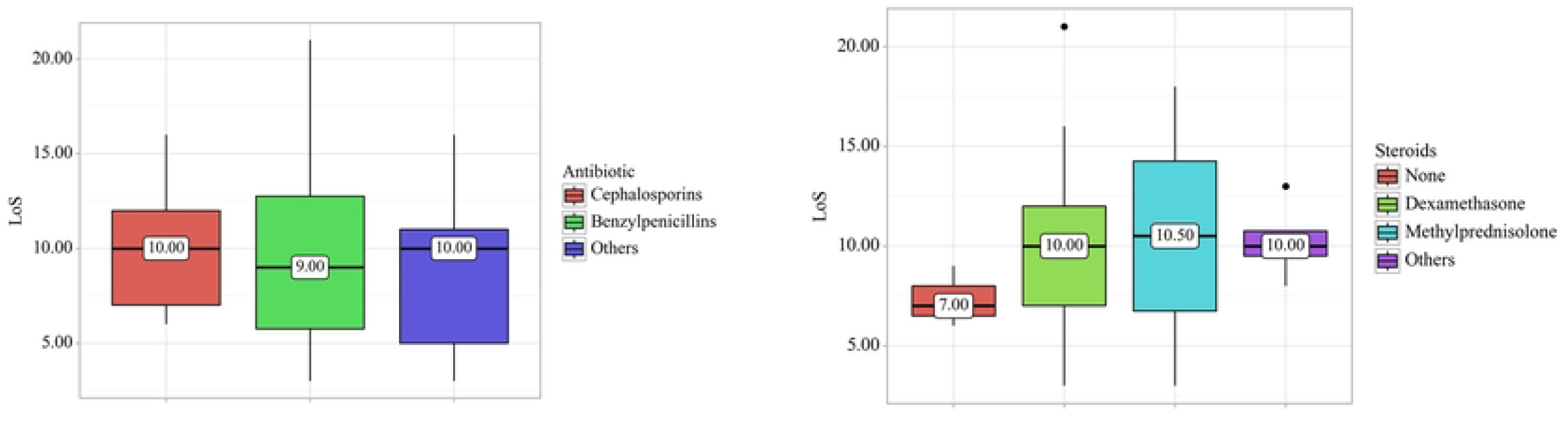
Box plots illustrating the Effect of steroids and antibiotics on Length of Stay (LoS). No significant changes were observed in patients receiving either steroids or antibiotics

### 3.4. Effect of laboratory variables on outcome

The non-survivor group had statistically higher serum creatinine levels on admission compared to the survivor group (137.30 [111.80 – 321.90] vs. 541.30 [506.10 – 570.60], p = 0.032) and GGT levels (75.60 [49.85 – 125.55] vs. 185.30 [163.75 – 442.35], p = 0.032). Total bilirubin levels were significantly higher in the non-survivor group compared to the survivor group (491.60 [343.95 – 563.30] vs. 25.90 [11.35 – 82.76], p = 0.010), as were direct bilirubin levels (257.10 [199.30 – 398.85] vs. 14.40 [5.84 – 53.80], p = 0.010). Additionally, urea levels were higher in the non-survivor group (30.80 [30.59 – 38.15] vs. 9.63 [8.20 – 21.25], p = 0.019).

Moreover, we conducted Receiver Operating Characteristic (ROC) analysis. We found that all variables had 100% sensitivity as potential predictors of death, except for GGT, which had a sensitivity of 66.8%. The specificity was lowest for GGT (77.1%), followed by serum creatinine (85.7%), total bilirubin and urea (both 88.6%), and the highest specificity was for direct bilirubin (91.4%) (Figure 4).

**Figure 4.**
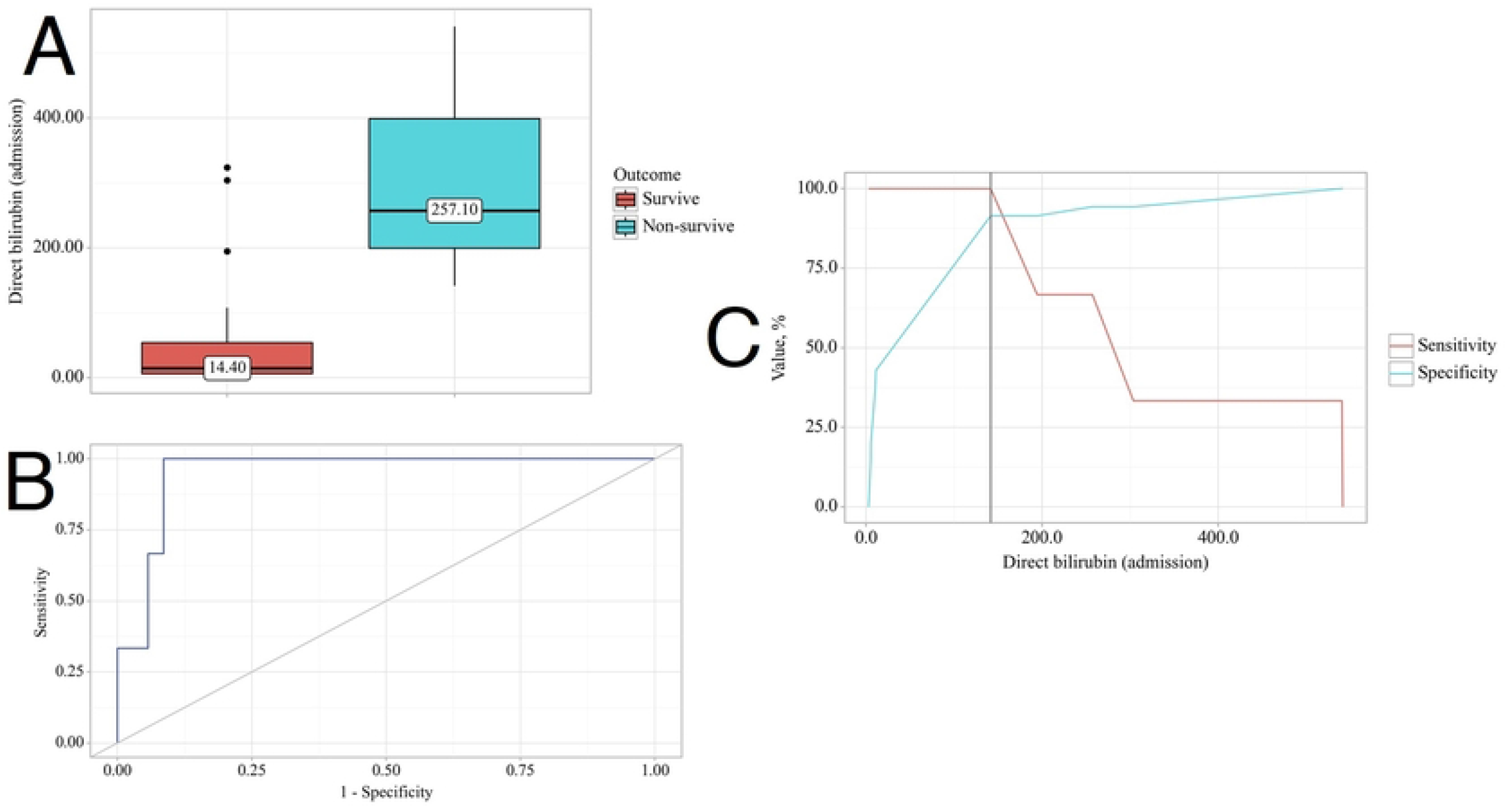
Direct bilirubin levels on admission as a potential predictor of lethality in leptospirosis. A. Box plot illustrating direct bilirubin levels on admission in surviving and non-surviving patients. B. ROC curve depicting the relationship between the probability of outcome and direct bilirubin. C. Analysis of sensitivity and specificity of outcome based on direct bilirubin levels.

## 4. Discussion

The Transcarpathian region experienced a significantly higher notification rate in 2023, with 150 out of 433 reported cases originating from this area. Situated in the western part of Ukraine, the region shares borders with Romania, Hungary, Poland, and Slovakia [8]. It sustains an above-average human population density, with 63% residing in rural regions, which may impact notification rates [18].

We observed that males had higher levels of transaminases such as AST, ALT, and GGT, but this did not affect outcomes. Our findings align with a study conducted in Germany, which revealed that male patients were more likely than female patients to be hospitalized and exhibit symptoms of severe leptospirosis, including jaundice, renal impairment, and haemorrhage [19].

Only four studies were found that investigated the role of steroids in clinical outcomes in leptospirosis. In three of these studies, methylprednisolone was administered at the initiation of treatment, albeit at varying doses. They suggested a potential beneficial role of steroids, particularly in patients with lung involvement [20-22]. However, the study that employed dexamethasone at the initiation failed to show a treatment benefit and, notably, reported an elevated incidence of nosocomial infections [23].

In regards to the effect of antibiotics on leptospirosis, we found a newly conducted meta-analysis involving 920 patients and 8 antibiotics. The analysis revealed that six antibiotics resulted in significantly shorter defervescence times compared to the control group. These antibiotics include cefotaxime, azithromycin, doxycycline, ceftriaxone, penicillin, and penicillin or ampicillin. However, antibiotics were not found to be effective in reducing mortality or hospital stays [24].

Lastly, concerning factors influencing leptospiral outcomes, we conducted our own meta-analysis of clinical predictors involving 1714 patients with leptospirosis. We found that patients with severe outcomes were more likely to experience dyspnea, oliguria, and hemorrhagic symptoms compared to non-severe patients [9].

## 5. Conclusions

We observed that dexamethasone had a statistically significant effect on certain clinical variables, such as LYM, MID, GRA, Platelet levels, AST, direct bilirubin, and urea levels. Similarly, cephalosporins showed significant effects on LYM, MID, Platelet, AST, total bilirubin, direct bilirubin, and urea levels. Notably, direct bilirubin levels emerged as one of the strongest predictors of death in leptospirosis, exhibiting high sensitivity and specificity. These findings have implications for clinicians not only in the Transcarpathian region but also for practitioners worldwide.

## Data Availability

The data presented in this study are available on request from the corresponding author.

## 8. Author Contributions

Conceptualization, and writing—original draft preparation, P.P.; writing—review and editing, M.P., A.K., V.O., I.P.; supervision, O.K.; visualization, O.K., P.P.; All authors have read and agreed to the published version of the manuscript.

## 9. Institutional Review Board Statement

The study was conducted in accordance with the Declaration of Helsinki and approved by the Ethics Committee of Uzhhorod National University (protocol code 5/1 on 30 May 2024).

## 10. Data Availability Statement

The data presented in this study are available on request from the corresponding author.

